# Reliability and criterion validity of a low-cost handgrip dynamometer: The Camry

**DOI:** 10.1101/2024.06.25.24309304

**Authors:** Lucía Sánchez-Aranda, Javier Fernández-Ortega, Isabel Martín-Fuentes, Ángel Toval, Gregor Jurak, Jonatan R Ruiz, Tamás Csányi, Francisco B. Ortega

## Abstract

**Background:** Handgrip strength has been related with multiple health outcomes, including all-cause mortality and morbidity. Handgrip testing is a highly valid and reliable method, included in evidence-based fitness test batteries from preschool to older ages. Previously, Jamar and TKK dynamometers have shown good reliability and validity against known weights. However, the cost of these dynamometers is the major limitation for implementing handgrip strength testing in certain countries and settings, as well as at large scale. Recently, a ten times cheaper model (Camry Dynamometer) has been used in fitness surveillance systems, though its reliability and validity, compared to known weights and other well-validated dynamometers, remains unknown. Therefore, the aims of the current study were to test to examine test-retest reliability, inter-model reliability (comparing a Camry dynamometer with 3000 uses versus a new Camry dynamometer), and inter-instrument reliability (Camry versus TKK dynamometer) of Camry dynamometer, using calibrated known weights.

**Methods:** A digital TKK 5401 dynamometer and two Camry EH101, a new and an “old” (3000 uses), dynamometers were used. Intra-instrument and inter-instrument reliability, and criterion related validity were assessed comparing the measures of the dynamometers with calibrated weights using the Bland and Altman’s method.

**Results:** Intra-instrument (retest minus test) reliability was very high (systematic error for test-retest reliability: New Camry = 0.01±0.49kg; Old Camry = -0.10±0.49kg; TKK = 0.14±0.77kg). The comparison between instruments showed small mean differences between Camry dynamometers and TKK (New Camry VS TKK: 0.84±0.79kg; Old Camry VS TKK: 0.88 ±0.85kg). The systematic error between New and Old Camry dynamometers was 0.03±0.57kg. Criterion-related validity showed smaller magnitude systematic errors in the Camry than TKK instruments (New Camry: -0.21±0.35kg; Old Camry: -0.18±0.79kg; TKK: -1.07±0.75kg).

**Conclusions:** Our results support that Camry dynamometer has an excellent reliability and validity and is therefore a more affordable alternative for handgrip strength assessment. Our results also indicate a good durability of these devices, and that calibration is not necessary, even after several thousands of uses.

## INTRODUCTION

Handgrip strength tests measure isometric grip strength, which has demonstrated to be a reliable indicator of whole-muscle strength^1,2^ and muscle mass^3^. Moreover, maximal handgrip strength (hereinafter just handgrip strength) has demonstrated to be an excellent biomarker of health in a wide range of populations^4–7^. The evidence has shown that handgrip strength is negatively associated with a wide set of pathological conditions such as cancer^8^, dementia^9^, cardiovascular disease^10^, or sarcopenia^11^. Consequently, the assessment of handgrip strength may be of importance from a clinical, therapeutic, occupational, public health and sport performance perspective. Due to this evidence handgrip has been widely used in research^3,9^. The assessment of handgrip strength is part of evidence-based fitness test batteries for different age groups, such as the PREFIT battery for children 3-5 year-old^12^, the ALPHA battery, for children 6-18 year-old^13^ and the ADULT-FIT battery, for adults 18-64 year-old^14^. The test is performed with hand dynamometers, which are easy and portable gadgets. Some handgrip dynamometers, such as the Jamar (Sammons Preston, Inc., Bolingbrook, IL, USA) and the TKK (Takei, Tokyo, Japan), have been widely used and have demonstrated to be reliable and valid for the assessment of handgrip strength^14^. Their reliability and validity have been tested using calibrated known weights, showing the TKK dynamometers a highest test-retest reliability and criterion-related validity^15,16^. However, the cost of these (Jamar and TKK) dynamometers has been relatively expensive (>400 euros), which might be a limitation for its use in certain settings, as well as in large-scale fitness monitoring (e.g. in school settings) and surveillance systems. Recently, a much cheaper version, the Camry EH101 dynamometer (cost roughly 40-50 euros), has been used in fitness surveillance systems, such as in Hungary and Slovenia, for large-scale nation-wide fitness assessment^17,18^. Furthermore, handgrip strength testing has been integrated into the FitBack platform^19^, offering a global opportunity to develop fitness monitoring systems. Consequently, the cost of dynamometers becomes a crucial factor in determining the feasibility of such systems, and the Camry EH101 is one of the low-cost alternatives. However, the reliability and validity against known weights has not been tested in the Camry EH101 dynamometer. In addition, testing the durability of the dynamometers is important as the measurement error could potentially increase after being heavily used for years^20^.

The main aim of the current study was to investigate the test-retest reliability and criterion related validity of the Camry EH101 dynamometer, using calibrated known weights. In addition, we compared an old (over 3000 uses for 8 years) versus a new (just bought and used for this study) Camry EH101 dynamometer to investigate whether the accuracy of the measurement change with the use and time. Finally, we also included the digital TKK dynamometer (model 5401, by Takei), for comparison purposes since it has previously shown to be highly reliable and valid^15,16^.

## METHODS

### Instruments

A digital TKK 5401 dynamometer (Takei, Tokyo, Japan) with a range of measurement from 5.0 kg to 100.0 kg, two Camry digital hand dynamometers EH101 (Sensun Weighing Apparatus Group Ltd, Guangdong, China; one new and one old with more than 3000 uses and 8 years in use in school settings) with a range of measurement from 0.0 kg to 90.0 kg (Figure 1), and known weights were used for this study. For the verification of the weights, we used a high precision SECA scale (Model 769; SECA, Hamburg, Germany). Dynamometers, weighs and the scale were calibrated by the manufacturer. Following previous studies in this field^15,16^, we assumed the validity of SECA scale as criterion method as we could not test the dynamometers against any other gold standard method for weight. However, we tested its test-retest reliability by assessing the inter-trial difference when measuring twice the weights in SECA scale, being 0.06±0.12 kg, indicative of high reliability.

**Figure 1.**
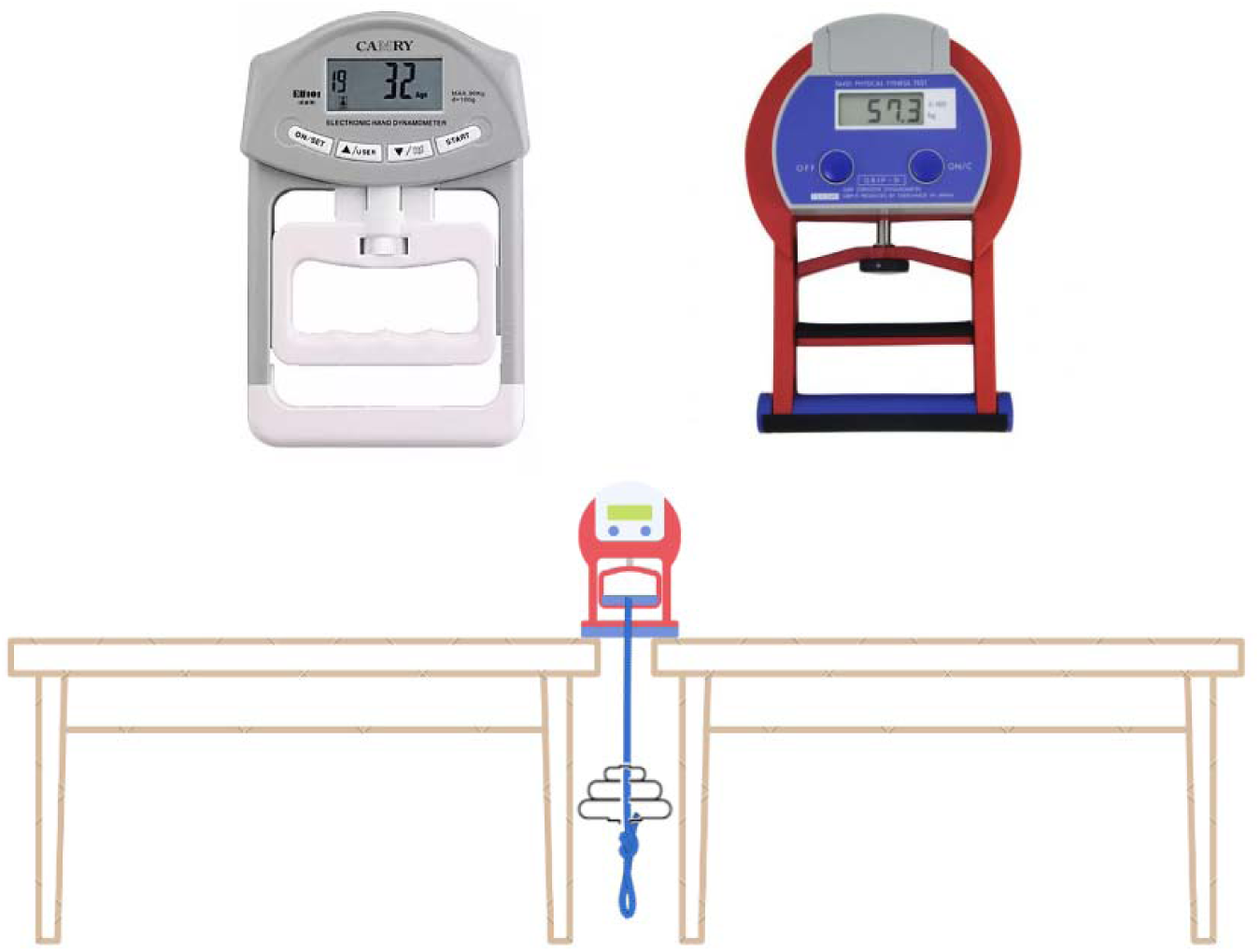
Camry EH101 (left) and TKK 5401 digital (right) dynamometers used in this study, and graphical illustration of the set-up for the measurement, showing the TKK 5401 as example.

### Procedures

The dynamometers were tested in a randomized order, using the known weights, ranging from 1 to 70 kg with increments of 1 kg up to 20 kg and increments of 5 kg up to 70 kg. The weights were also used in a randomized order. In total, 30 weights measurements were taken twice (test-retest) with each dynamometer. Dynamometers were placed on two tables. The weights were suspended from the dynamometer with a loading belt, hanging between the tables (Figure 1). The center of the dynamometers handle was previously marked with tape for the constant placement of the loading and 5 cm grip span was used for all the measurements, since this grip span falls within the range of optimal span observed in men and women^21^.

### Statistical analysis

We applied the Bland-Altman’s method^22^ to investigate intra-instrument (test-retest) and inter-instrument (comparing the three dynamometers New and Old Camry EH101, and TKK 5401) reliability, and criterion-related validity (comparing dynamometers with known weights). Mean difference (systematic bias) between measurements and 95% limits of agreement (mean difference ± 1.96 of standard deviation of the difference) were calculated. Bland-Altman plots were created to visually represent individual variation of the measurements in the relationship between the measurement’s differences and means^22^.

Additionally, heteroscedasticity, considered as non-consistency of error among weights increments, was calculated as the comparison of the measurement’s differences between weights ≤15 kg and >15kg. We used this grouping to simulate populations with low handgrip performance such as elderly, diseased people or very young children, and test whether the reliability and validity would differ between low and medium-high performers. For heteroscedasticity analyses, the differences were transformed to absolute values (i.e. multiplying the negative values by -1) and compared by using one-way ANOVA, with the absolute differences as dependent variable and the weight groups (≤15 kg and >15kg) as the fixed factor. Significant differences, expressed as *p*<0.05, would confirm heteroscedasticity. Additionally, visual inspection of the Bland-Altman plots can inform on whether there is or not heteroscedasticity. All statistical analyses were conducted using R software version 4.3.1.

## RESULTS

### Reliability

Table 1 shows the mean differences among repeated measures with the same instrument (intra-instrument reliability) and with different instruments (inter-instrument reliability). According to the intra-instrument test-retest reliability, New Camry dynamometer had the smallest mean error (0.01±0.49kg), followed by its old version (-0.10±0.49kg) and TKK dynamometer (0.14±0.77kg). When comparing between instruments, the mean differences between the two Camry dynamometers (Old vs. New) resulted smaller (0.03±0.57kg) than the differences between Camry dynamometers and TKK (New Camry vs. TKK: 0.84±0.79kg; Old Camry vs. TKK: 0.88 ±0.85kg) (Table 1). Heteroscedasticity using absolute differences was tested to investigate whether the variability between trials (test-retest) or between instruments changed as the magnitude increased, i.e. comparing weights ≤15 and >15 kg. Overall, heteroscedasticity was present in all comparisons, being significant (P<0.05) in most of them, indicating that the variability is larger (i.e. the reliability is lower) at higher weights (Table 1). However, the errors were small (i.e. <0.5kg differences between higher and lower weights). The intra-instrument and inter-instrument reliability analyses are graphically represented in Figures 2 and 3 using Bland–Altman plots.

**Figure 2.**
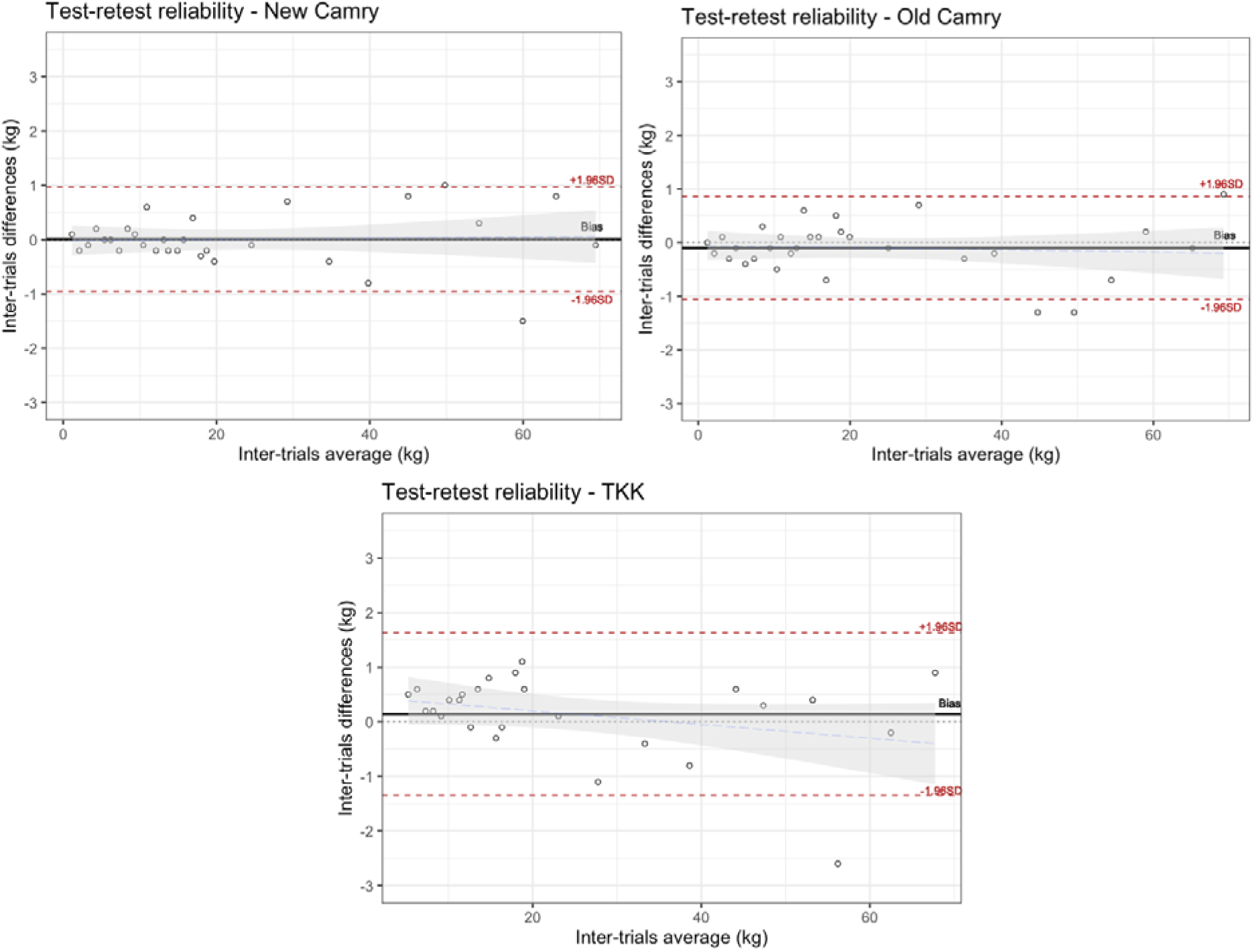
Bland-Altman plots showing the test-retest reliability (retest minus test).

**Figure 3.**
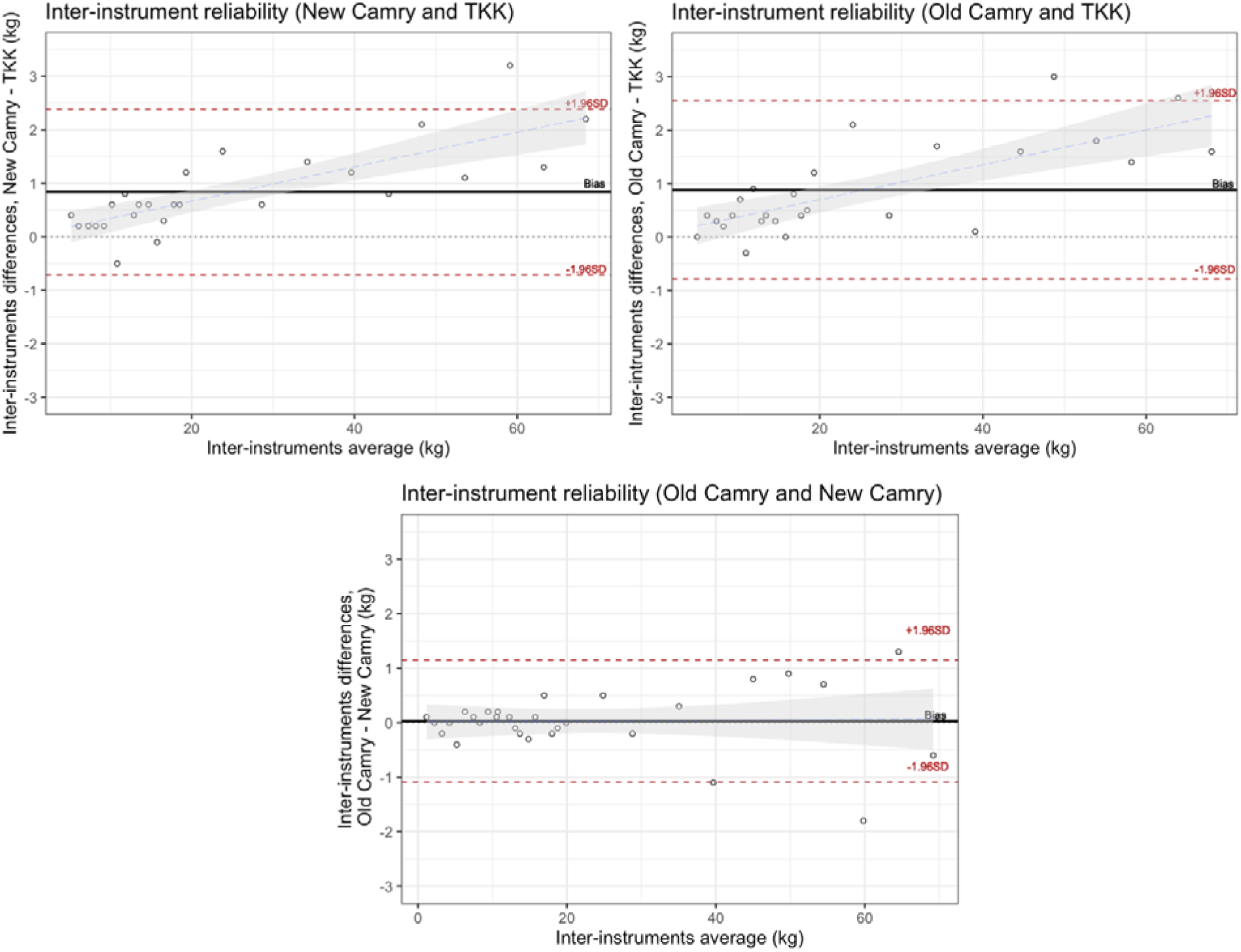
Bland-Altman plots showing the inter-instrument reliability.

**Table 1.** Intra-instrument and inter-instrument reliability of the Camry and TKK dynamometers.

| Dynamometer | All weights<br>(kg) | Weights <15 kg<br>M ± SD | Weights >15 kg<br>M ± SD | Difference<br>between weights<br>p<sup>a</sup> |
| --- | --- | --- | --- | --- |
|  | M ± SD | (Absolute<br>values*) | (Absolute<br>values*) |  |
| Comparison using the same instrument (intra-instrument reliability: retest minus test) |  |  |  |  |
| New Camry EH101 | 0.01±0.49 | 0.16 ±0.15 | 0.52±0.41 | 0.005 |
| Old Camry EH101 | -0.10±0.49 | 0.23 ±0.17 | 0.49±0.42 | 0.036 |
| TKK Model 5401 | 0.14±0.76 | 0.40±0.23 | 0.69±0.63 | 0.112 |
| Comparison between instruments (inter-instrument reliability) |  |  |  |  |
| New Camry EH101 minus<br>TKK | 0.84 ± 0.79 | 0.43±0.21 | 1.22 ±0.81 | 0.002 |
| Old Camry EH101 minus<br>TKK | 0.88 ±0.85 | 0.38±0.24 | 1.28 ±0.90 | 0.002 |
| Old Camry EH101 minus<br>New Camry EH101 | 0.03 ±0.57 | 0.15 ±0.11 | 0.61 ±0.51 | 0.004 |

All means (M) and standard deviations (SD) represent the difference between trials or instruments.
\* The analysis split by weights groups was conducted transforming the real difference variable into absolute differences by multiplying all negative values by -1, so that a higher positive value is indicative of higher variation between test and retest, or between instruments, in any direction.
<sup>a</sup> Analysis of covariance (ANOVA) was performed with the absolute differences as dependent variable and weight group as fixed factor.

### Validity

Criterion-related validity results (dynamometers measures against known weights) are presented in Table 2. Criterion-related validity showed a small-magnitude negative systematic error for New Camry (-0.21±0.35kg) and Old Camry (-0.18±0.79kg) dynamometers. This error was found to be slightly larger for TKK dynamometer (-1.07±0.75kg). The heteroscedasticity analysis showed a significant increment on the absolute mean difference for weighs above 15 kg, as compared with lighter weights (≤15kg), with the largest difference observed in the case of TKK dynamometer (roughly 1kg larger error in heavier weights) and small differences for the Camry dynamometers (<0.5kg). Results from criterion-related validity were additionally plotted using Bland-Altman’s method for each of the dynamometers studied (Figure 4). While there was no obvious association between the real differences and the magnitude between the Camry dynamometers and the known weights, there was a clear negative association in the case of the TKK dynamometer, indicating that the larger is the magnitude (i.e. higher weights) the larger was the underestimation of the TKK dynamometer compared to known weights.

**Figure 4.**
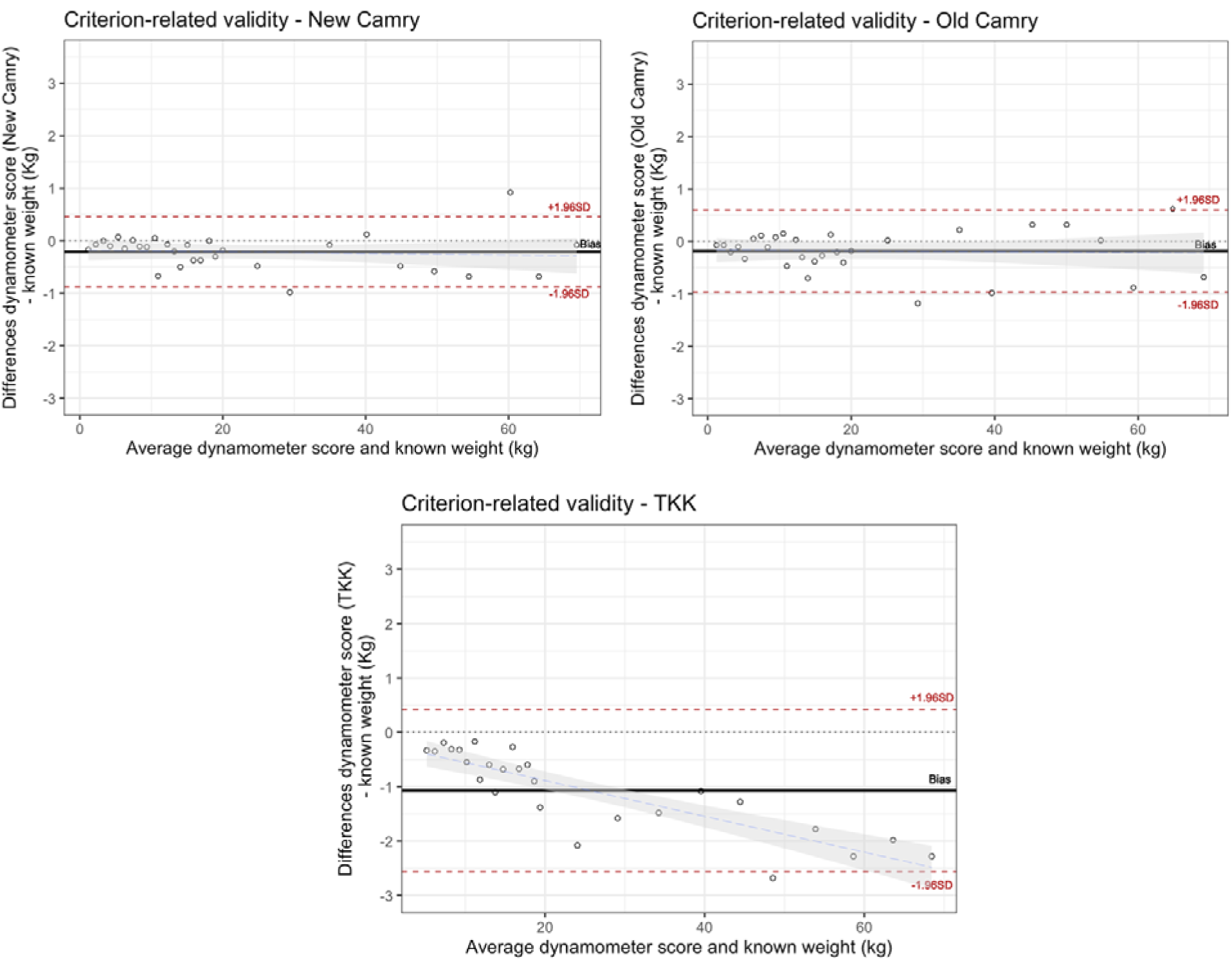
Bland-Altman plots showing the criterion validity of the TKK digital dynamometer and the Camry EH101 dynamometers against known weights.

**Table 2.** Criterion related validity of the Digital TKK and Camry dynamometers compared against known weights.

| **Dynamometer** | **All weights** | **Weights <15 kg,**<br>**M ± SD** | **Weights >15 kg,**<br>**M ± SD** | **Difference**<br>**between**<br>**weights, p<sup>a</sup>** |
| --- | --- | --- | --- | --- |
|  | **M ± SD** | **(Absolute**<br>**values\*)** | **(Absolute**<br>**values\*)** |  |
| **New Camry EH101** | -0.21±0.35 | 0.16±0.18 | 0.42±0.31 | 0.009 |
| **Old Camry EH101** | -0.18±0.79 | 0.21±0.19 | 0.43 ±0.36 | 0.05 |
| **TKK Model 5401** | -1.07±0.75 | 0.50±0.29 | 1.49 ±0.70 | <0.001 |

All means (M) and standard deviations (SD) represent the difference of dynamometers' values minus known weights, so that a negative value is indicating that the dynamometer is underestimating the known weights and viceversa.
\* The analysis split by weights groups was conducted transforming the real difference variable into absolute differences by multiplying all negative values by -1, so that a higher positive value is indicative of higher variation between the dynamometer and known weights in any direction.
<sup>a</sup> Analysis of covariance (ANOVA) was performed with the absolute differences as dependent variable and weight group as fixed factor.

## DISCUSSION

Our study contributes to the existing knowledge by providing novel findings about the objectively assessed (against known weights) reliability and validity of a low-cost handgrip dynamometer, the Camry dynamometer. First, test–retest reliability was excellent (i.e., mean error ≤0.1kg) for all the dynamometers analyzed, being slightly superior for New and Old Camry dynamometers (i.e. lower systematic and random errors) than for the TKK dynamometer. Second, the comparison between the New and Old Camry showed strong consistency, indicating a good durability. However, the comparability was lower between the Camry dynamometers and the TKK, particularly at high weights. Third, the New and Old Camry dynamometers showed a high validity against known weight, with a mean difference (bias) of -0.2kg, and a random error (limits of agreement) slightly smaller than 1kg. On the other hand, the TKK showed on average roughly 1kg underestimation compared to known weights, progressive increasingly underestimation as the weights increased, suggesting it would have a larger error in individuals with high handgrip strength.

Known weights have been previously used for the assessment of reliability and validity in different dynamometers^15,16,23,24^. However, to our knowledge, no previous study has targeted the reliability and criterion related validity of Camry dynamometer using known weights, which has important implications due to its markedly lower cost. When testing reliability and validity in different population groups, it is important to consider the inherent variability of human biology when performing repeated measurements. Our results from the test-retest reliability analysis demonstrate the excellent reliability of Camry dynamometer, when using known weights. We found a systematic error between trials of 0.01±0.49 kg for the New Camry dynamometer and -0.10±0.49 kg for the Old one. These results are in line with Latorre Román et al.^25^ findings, who analyzed test-retest reliability Camry EH101 dynamometer in a healthy preschool children population (n=1215; mean age = 4.32 ± 1.05 years) and reported similar differences between trials (0.11±0.69 kg) with an Intraclass Correlation Coefficient (ICC) (being a value of 1 known as perfect reliability), of 0.969. Similarly, Mani et al.^26^ reported an ICC of 0.95, when testing Camry EH101 reliability in 114 healthy adults, and Cao et al.^27^ found also a highly stable ICC (0.737) among 599 female college students (18.7 ± 1.00 years). In the case of TKK digital dynamometer, we found a systematic error of 0.14±0.76 kg between test-retest measurements, which can also be considered good reliability and is in line with previous results from España-Romero et al.^15^, who found test-retest systematic error of 0.02kg for TKK 5101 dynamometer, and Cadenas-Sánchez et al.^16^, who reported intra-instrument systematics errors for several TKK models ranging from 0.09±0.65 to -0.33±0.69. However, the variability of the measure changed slightly as the weights increased, suggesting its reliability might differ slightly between people with lower and higher handgrip performance.

Comparability among different dynamometers is crucial for interpreting and pooling data from different studies. To our knowledge, this is the first study comparing Camry and TKK digital dynamometer. TKK digital dynamometer has previously been validated, even showing lower systematic error against the Jamar dynamometer^15^, widely and classically used. We found that Camry dynamometer might show slightly higher handgrip strength values as compared with TKK digital dynamometer. In contrast, a previous study in a geriatric setting (n=1064; mean age= 66=±=7.7 years old)^28^ showed lower values in the Camry EH101 when comparing to Jamar dynamometer, being this difference 0.5 kg in men and 0.6 kg women. Similar results were found by Andrade et al.^29^ who also concluded that the Camry EH101 dynamometer showed lower grip strength, with an average difference of -0.11 kg for the right hand and -0.30 kg for the left hand, in 220 older adults (73.1 ± 6.3 years old). Additionally, we assessed the durability of the Camry dynamometer by comparing a new device with an old one. Our findings revealed a robust durability, with a systematic error of 0.03±0.57 kg, which suggest there is no need to calibrate the device after many uses, at least up to the roughly 3000 uses over an 8-year period of the dynamometer used in this study.

According to criterion-related validity (comparison with known weights), our analysis revealed a small-magnitude negative systematic error for the three dynamometers. The New and Old Camry dynamometers showed better agreements with the known weights, with a systematic error of -0.21±0.35kg and -0.18±0.79kg respectively, whereas TKK digital dynamometer presented greater underestimation, with a systematic error of - 1.07±0.75kg and a tendency to increase the error more markedly as the magnitude of the measure increases (i.e. heteroscedasticity). These findings are in concordance with those described by España-Romero et al.^15^ and Cadenas-Sánchez et al.^16^ whose studies showed heteroscedasticity as well among TKK digital dynamometers. Likewise, these previous studies also found the Jamar, TKK and DynEx dynamometers underestimated between 0.5 and 2.6kg compared to known weights^16^. Thus, Camry dynamometers seem to have similar and even better agreement with known weights compared to other dynamometers.

The main strength of the present study is that we determined the reliability and validity of the low-cost dynamometer Camry EH101 using known weighs, which avoid the human variability. Future studies should replicate our findings and include other low-cost dynamometers, as well as other well-known dynamometers (e.g. Jamar) for comparability purposes.

### Conclusion

In conclusion, the findings indicate that the Camry EH101 dynamometer has an excellent reliability and validity and can be therefore used for handgrip strength assessment in different populations with higher or lower handgrip strength levels. Our results suggest also that these properties remain after being heavily used for several years, suggesting these devices are durable and do not need re-calibration. Most importantly, due to its reduced cost, it seems an excellent value-for-money alternative for the assessment of handgrip strength in large scale population studies or for monitoring and surveillance systems, as well as for individual end-users in clinical, educational and sport settings.

## Data Availability

All data produced in the present study are available upon reasonable request to the authors

## DECLARATIONS

The authors declare no competing interest.

